# Tuberculosis and depression: cultural dynamics of comorbidity among Pashtun communities in Pakistan and Afghan refugees

**DOI:** 10.64898/2026.02.12.26346137

**Authors:** Fayaz Ahmad, Fatima Khalid, Zohaib Khan, Noor Sanauddin, Maryiam Rahim, Sikandar Sultan, Shaista Rasool, Mirrat Gul, Farooq Naeem, Firaz Khan, Nishani Fonseka, Abbie Milner, Saima Sheikh, Saeed Farooq

**Affiliations:** Institute of Public Health & Social Sciences, Khyber Medical University, Peshawar, Pakistan; Office of Research Innovation and Commercialization, Khyber Medical University, Peshawar, Pakistan; Department of Sociology, University of Peshawar, Peshawar, Pakistan; Department of Psychiatry and Behavioral Sciences, Mayo Hospital, Lahore, Pakistan; University of Toronto & Centre for Addiction & Mental Health, Toronto, Canada; Institute of Mental Health and Behavioral Sciences, Khyber Medical University.; School of Medicine, Keele University, Staffordshire, UK

**Keywords:** Tuberculosis (TB), depression, comorbidity, cultural perceptions, Pakistan, Pashtun community, Afghan refugees

## Abstract

**Objectives:** Tuberculosis (TB) remains a significant global health challenge, ranking among the top ten causes of death worldwide. Pakistan is the fifth-highest burden country for TB globally. Patients with TB often develop depression, where cultural factors play a profound role.

This study explores the cultural perceptions and beliefs that influence TB-depression comorbidity among the Pashtun ethnic population of Pakistan and Afghan refugees, with a broader aim of developing a culturally appropriate intervention for the treatment of depression in TB.

**Study design:** We conducted a qualitative study across TB treatment centres in the Haripur and Peshawar districts of Khyber Pakhtunkhwa, Pakistan.

**Methods:** The study comprised 29 in-depth interviews and 11 focus group discussions, involving 101 participants across three groups: healthcare providers, TB patients, and carers. We analyzed the data using a deductive-inductive thematic analysis approach and employed the Southampton framework for cultural adaptation of interventions.

**Results:** We identified four central themes. The first theme described the cultural norms of the region, which influence physical and psychological conditions related to TB and depression. The second theme highlighted the role of the belief system in relation to the TB-depression comorbidity. The third theme explained the existing stigma associated with TB and depression. The last theme informed the development of a culturally appropriate psychotherapy-based intervention.

**Conclusion:** Gender disparities, social stigma, family dynamics, lack of social support, and belief systems are culturally driven issues linked to TB-depression comorbidity. Psychosocial interventions must consider and integrate the socio-cultural context of the target population.

## Introduction

Globally, tuberculosis (TB) is among the top ten causes of death, with an estimated 10.6 million cases of active TB documented in the year 2021, and 1.6 million deaths due to tuberculosis ^1^. More than 95% of TB-related fatalities have been reported in low- and middle-income countries (LMICs) ^2^, with Pakistan ranking fifth among these TB high-burden countries globally ^3^. Pakistan bears almost two-thirds of the TB burden in the Eastern Mediterranean Region (EMRO), with an estimated half a million new cases of drug-susceptible TB and 15,000 new cases of drug-resistant TB annually ^3^.

Tuberculosis patients often develop symptoms of depression, including low mood, emptiness, hopelessness, and suicidal thoughts ^4,5^. The bidirectional relationship between TB and depression affects a patient’s quality of life, leads to suboptimal adherence to anti-TB treatment, which can advance to drug-resistant TB, and ultimately results in a higher risk of mortality ^6^. Individuals are more susceptible to infectious disease conditions like TB or HIV due to their low levels of mental health ^7,8^. Community-based data from 48 LMICs reports a 23.7% prevalence of depression in TB patients ^7^. This number is almost doubled in Pakistan, where 42.8% of TB patients were suffering from depression ^9^.

TB and depression are two prominent health challenges for LMICs. Both conditions share a range of fundamental social determinants, such as a lack of health-related knowledge and understanding, the impact of cultural norms, social class differences, and challenges to healthcare infrastructure, further aggravated by the absence of social protection ^2^. Cultural factors play a significant role in the experience and conceptualisation of mental health issues ^10,11^. These cultural perspectives and formal biases against specific groups within communities further contribute to the overall burden of illness in these population subgroups ^12^. It has been well established that TB and mental health stigma are significant barriers to seeking and receiving care in LMICs, and can result in multiple social and functional consequences throughout both conditions ^13^.

The stigma related to TB may cause low mood and depression. A patient may feel a sense of shame and guilt in response to the community’s negative attitudes and behaviours toward them, promoting discrimination, social seclusion, and ultimately depression ^14^. Similarly, a diagnosis of TB is often accompanied by a fear of society’s rejection, resulting in loss of self-respect, fear of infecting loved ones, and a general role limitation ^15^. The psychosocial aspects of TB and depression are particularly pertinent in LMIC contexts, where inadequate health systems must rely on social support to address disease conditions ^15^. Identification of these social and cultural drivers will play a vital role in mental health recovery and service delivery ^16^.

Cognitive Behavioural Therapy (CBT) has proven effective for mental health disorders such as depression and anxiety ^17^. To develop a CBT-based intervention for a specific population, it is essential to thoroughly understand their socio-cultural and religious values ^18^. Interventions adapted to particular populations are four times more effective than more general interventions^19^. It is, therefore, essential to explore the complex social and cultural factors of the region that influence the management of TB and depression in LMICs, to inform a more patient-centric approach to the treatment of TB-depression comorbidity.

This study was conducted in the Khyber Pakhtunkhwa (KP) province of Pakistan. Participants included Pakistani nationals as well as Afghan refugees, as KP has become home to almost three million refugees since the ongoing conflict in Afghanistan. Afghanistan is also a TB high-burden country among the EMRO nations, with an incidence of 189 per 100,000 population ^20^. This study aimed to thoroughly explore Pashtun culture and its influence on patients’ access, engagement, and treatment adherence. The broader aim was to inform the development of a CBT-based therapy manual for the treatment of depression in TB patients. This is a unique study from Pakistan, which specifically targets the TB-depression comorbidity, combining the perceptions of both service providers and service users (including carers).

## Materials and methods

We conducted a qualitative study, employing the Southampton adaptation framework ^21^, to explore the Pashtun cultural factors that should be addressed for the adaptation of a CBT therapy. The Southampton framework is deemed suitable for this study as it is based on systematic observations conducted in a developing country and can identify themes sensitive to mental health issues within the local cultural context of KP.

### Study settings

The National TB Control Program (NTP) of Pakistan provides services for both drug-resistant TB (DRTB) and drug-sensitive TB (DSTB) through DOTS (Directly Observed Treatment Short Course) centers located in every district of Pakistan, also referred to as basic management units (BMUs). This study was conducted in the BMUs providing DSTB services, located within primary, secondary, and tertiary healthcare facilities in two districts of Khyber Pakhtunkhwa Province, Pakistan: Peshawar and Haripur. The two districts were selected purposively based on the high TB burden, to capture both the Pakistan and Afghan Pashtun communities, and to achieve a mix across the three healthcare levels.

### Participants and recruitment

We purposively recruited three groups of participants for each district: a) TB patients, b) carers of the patients, and c) healthcare providers. We conducted a total of 29 in-depth interviews (IDIs) and 11 focus group discussions (FGDs), each comprising 6-8 participants. In Peshawar, we conducted 16 IDIs and 7 FGDs, while in Haripur, we conducted 13 IDIs and 4 FGDs. To ensure a maximum variation in the sampling, we recruited participants of both genders and from both nationalities (Pakistani and Afghan), aged 20-64 years. The research team identified eligible participants with the help of DOTS facilitators.

### Inclusion criteria

All patients with TB who had completed at least one month of their treatment were recruited. Similarly, any caregiver involved in the care of and escorting TB patients to the BMU was eligible for recruitment. Healthcare providers included both DOTS facilitators and medical officers (MOs), as every BMU comprises at least one person in each position. The DOTS facilitators maintain a close emotional connection with patients, providing counseling and administering medications, while the MOs mainly contribute from a clinical standpoint.

### Data collection

We collected data from January to March 2023 to gather information about the subjective experiences related to various aspects of TB care and how culture may have influenced those experiences. Specifically, we inquired about treatment adherence, cultural barriers to treatment, and mental health conditions. We developed separate topic guides for each participant group, which were translated into local languages, Urdu and Pashto. The initial versions of the topic guides were pilot-tested in the field and later refined based on the pilot results. Each FGD lasted from 50 to 70 minutes, while IDIs took between 30 to 45 minutes. Participants were offered an opportunity cost in the form of a cash payment (2000 PKR, equivalent to £6).

IDIs and FGDs were conducted by a research team trained in qualitative research methods and qualitative data collection. Written informed consent was obtained from all participants to take part and to be audio-recorded. All the interviews were transcribed verbatim, translated into English, anonymised, and proofread against the audio recordings by the research team.

### Data analysis

We employed a framework analysis approach, utilizing a combination of deductive and inductive techniques. First, we developed a thematic framework based on the research questions, topic guides, thematic analysis of five transcripts, and the Southampton framework. The data were then subjected to the thematic framework with a focus on exploring the socio-cultural and religious factors involved in the interplay between TB and depression. The thematic framework was systematically applied to the transcribed data, including summaries of participants’ responses and verbatim quotes. The charted data were reviewed and analyzed to compare views, seeking patterns, connections, and explanations. After several iterations and cross-checks of data among the analysis team members, summary findings were finalized for each participant group. Finally, we triangulated the key findings from the three participant groups, examining their similarities and differences.

## Results

We recruited 101 participants, with 29 in IDIs and 72 in FGDs (Table 1). Four central themes were identified, describing the cultural influences related to depression among TB patients. The following sections elaborate on these themes.

**Table 1:** Socio-demographic characteristics of participants.

| Characteristics |  | Peshawar |  |  |  | Haripur |  |  |  |
| --- | --- | --- | --- | --- | --- | --- | --- | --- | --- |
|  |  | FGDs |  | IDIs |  | FGDs |  | IDIs |  |
|  |  | Male | Female | Male | Female | Male | Female | Male | Female |
| No. of participants |  | 33 | 13 | 9 | 7 | 12 | 14 | 6 | 7 |
| Age Mean (range) |  | 43.68<br>(22-64) | 42.8<br>(24-62) | 45.4<br>(34-63) | 39.6<br>(23-58) | 43.3<br>(20-63) | 38.96<br>(24-59) | 39.3<br>(32-56) | 47<br>(33-60) |
| Nationality of the participants | Pakistani | 20 | 13 | 5 | 3 | 6 | 14 | 4 | 6 |
|  | Afghan | 13 | 0 | 4 | 4 | 6 | 0 | 2 | 1 |
| Marital status | Married | 29 | 10 | 9 | 5 | 8 | 12 | 5 | 7 |
|  | Unmarried | 4 | 3 | 0 | 2 | 4 | 2 | 1 | 0 |
| Type of participant | Patient | 15 | 6 | 3 | 4 | 12 | 6 | 3 | 2 |
|  | Carer | 13 | 6 | 3 | 3 | 0 | 8 | 2 | 2 |
|  | Health professional | 5 | 1 | 3 | 0 | 0 | 0 | 1 | 3 |
| Employment status | Employed | 30 | 2 | 7 | 0 | 8 | 3 | 6 | 4 |
|  | Unemployed | 3 | 11 | 2 | 7 | 4 | 11 | 0 | 3 |
| Level of education | No formal education | 20 | 10 | 4 | 5 | 6 | 7 | 2 | 3 |
|  | Primary | 6 | 2 | 2 | 2 | 3 | 4 | 2 | 0 |
|  | Secondary & above | 7 | 1 | 3 | 0 | 3 | 3 | 2 | 4 |

### The role of socio-cultural factors

Overall, healthcare providers, TB patients, and carers shared a clear consensus that culture can seriously influence the treatment of TB and depression.

Healthcare providers elaborated that being female in Pashtun culture itself is a significant contributing factor in causing depression. They revealed that it is customary for females to be accompanied by an attendee (carer) to the hospital; it is difficult for females to seek care alone and to make decisions on their own about their health, continuation of treatment, and following instructions. Also, among Pashtuns, there is a belief that women often use illness as an excuse to avoid household chores or to seek the attention of family members. Here, at times, husbands take their wives to their parents’ houses when they are ill and bring them back when they are fully recovered. Moreover, Pashtun females are often subjugated, at times physically, which adds to their distress and culminates in mental health problems (Table-2). According to healthcare providers, Afghan TB patients frequently relocate between cities in search of employment, resulting in interruptions to their treatment.

**Table 2:** Views of healthcare providers.

| Theme | Respective Quote |
| --- | --- |
| **The role of socio-cultural factors** | "We have seen some Pashtun families who do not allow their females to go out of their homes even if they have a serious problem. We have seen patients die during delivery, but their families do not allow them to go outside for treatment in the absence of a male escort." (*Healthcare Provider 003-IDI-Peshawar*)<br>"According to her husband, she is making excuses and is completely all right. And her mother says that he [the husband] doesn't come with her to the TB center, and until she is completely cured, she will stay with me." (*Healthcare Provider 007-FGD-Peshawar*)<br>"In our Pashtun culture, inflicting suffering on females is also common. This is another factor. On one hand, the patient is ill, and on the other hand, she is facing hardship. This can lead to feelings of depression." (*Health Professional 002- IDI-Peshawar*) |
| **The role of the belief system and social support** | " We just treat them with little conversation by telling them to offer their prayers through which the depression will go away " (*Health Professional 004- IDI-Male-Haripur*) |
| **TB and depression: the dual stigma** | "Many girls who are engaged and getting married frequently ask us when the treatment will finish. They usually delay their ceremony, or in some cases, they hide their disease from their in-laws. There is poor acceptance in our culture." (*Health professional 001-FGD-Male-Peshawar*)<br>"Overall, in our community, this is a social stigma. Even highly qualified people have a negative attitude toward visiting psychiatrists and psychologists. We are humans, but in our society, we do not seek mental health treatment." (*Health professional 002-FGD-Male-Peshawar*) |
| **Culturally preferred and acceptable therapy** | "It should be provided at a facility rather than at home because it will lose its worth, just as the polio campaign. It should be bound to the facility and educate them that it will cure their TB and depression. (*Health Professionals 001-IDI-Female-Haripur*)<br>"Also, if we are provided with some training (tools and skills), then definitely, that would be better for us, and we will be able to counsel the patients of depression." (*Health Professionals 004-FGD-Male-Peshawar*)<br>"Because in a group people are not comfortable in sharing their problems and issues in front of others. It should be conducted individually; people hide their disease due to social stigma." (*Health professional 007-FGD-Male-Peshawar*) |

Reasons for depression shared by patients and carers were typically related to TB itself: most of the patients lived in a joint family system and were worried about the transmission of infection to other family members. Participants explained that many TB patients get disheartened, lose hope about their recovery, and end up suffering from depression, and such patients tend to avoid social gatherings (Table-3&4). Patients explained that for men, the notion of being the sole earner exacerbates the distress associated with the disease (Table-4).

**Table 3:** Views of carers.

| **Theme** | **Respective Quote** |
| --- | --- |
| **The role of socio-cultural factors** | "If a person takes the disease [TB] seriously, he/she gets half depressed already, and it gradually increases. If you don't take it seriously, there will be no depression." (*Carer 004-FGD-Male-Haripur*) |
| **The role of the belief system and social support** | "No matter how much you tell a patient you are well, it's the doctor who is treating him or her; if he says, Don't worry, you'll be all right, ' the patient will then improve. A doctor is the key to a patient's emotional well-being and recovery." (*Carer 002-FGD-Male-Peshawar*)<br>"Before coming to the TB facility, I took my mother to 'Hakim Kirdaani', someone recommended to my mother that she would get healed from illness if she got treatment from that hakim." (*Afghan carer 004-FGD-Male-Peshawar*) |
| **TB and depression: the dual stigma** | "Sadness is obvious because when your family members start keeping their distance from you, you feel unwanted and deprived." (*Carer 006-FGD-Male-Peshawar*)<br>"Ladies of villages make something out of the ordinary, which causes depression in our patients. You cannot express your illness in local communities." (*Carer 005-FGD-Male-Peshawar*) |
| **Culturally preferred and acceptable therapy** | "The healthcare provider knows them [patients] very well. Patients will be mentally relaxed with them. After counseling, the patient will be pleased and will have a good life, and their mind will be diverted from the TB." (*Carer 005- IDI- Male-Peshawar*)<br>"Counseling is even better than medicines; it is necessary to give counseling to TB patients periodically to give them strength/ courage that they are fine and will be cured soon, Insha' Allah." (*Afghan Carer 005-IDI-Male-Peshawar*) |

**Table 4:** Views of patients.

| **Theme** | **Respective Quote** |
| --- | --- |
| **The role of socio-cultural factors** | "I was in a bad condition; I couldn't even eat. I would hate everything, even normal conversation. Everything was unbearable." (*Patient 002-FGD-Female-Peshawar*)<br>"There are so many issues, you know. I have to take care of my family, for that I have to work, and then this disease [TB]....." (*Patient 003-FGD-Male-Haripur*) |
| **The role of the belief system and social support** | "I was visiting local doctors, but I wasn't recovering, which was extremely worrying. When TB was diagnosed, it was again sad news for me, but after visiting the TB center and now that I'm recovering from the disease, this gives me hope." (*Afghan Patient 008-FGD-Male-Peshawar*)<br>"They [healthcare providers] listen to us. They talk with sincerity and morality and have not shown any bad manners. The staff and environment are good; there are chairs where you can sit until your turn. May Almighty keep them happy." (*Patient 007- IDI-Male-Haripur*)<br>"Thank God, I got support from my family and friends. My family supported me emotionally and financially; they all took care of me. My mother used to give me medicines regularly with her own hands." (*Patient 005-IDI-Male-Haripur*)<br>"In our Afghan society, getting TB is a hazardous thing. The patient's routine utensils, bedding, and all other personal items will be kept separate. People wouldn't go near them [TB patients] because there was no proper treatment available, but in Pakistan, TB was treated." (*Afghan patient 005-FGD-Male-Haripur*)<br>"The treatment for depression is to pray five times a day and worship Allah. This is the only way to treat depression." (*Patient 011-IDI-Female-Peshawar*) |
| **TB and depression: the dual stigma** | "I regret that I was creating problems for people. I avoided interactions and conversation often with my family." (*Afghan patient 002-IDI-Male-Haripur*)<br>"I don't know, but when I look at myself, I want to cry. People don't even know about my illness; still, they ask why my color is becoming dull day by day. So, I wonder what they will say if they find out that I have TB." (*Patient 001-FGD-Female-Haripur*) |
| **Culturally preferred and acceptable therapy** | "For this [therapy], some books are also required so that patients remain busy with that. Books guide the TB patient and keep the patient's mind diverted." (*Patient 006-FGD-Male-Peshawar*) |

### The role of the belief system and social support

Many patients expressed that before receiving a diagnosis, they felt quite depressed. However, once they learned they had TB, they felt relieved, knowing it is curable and that treatment is provided free of charge. Conversely, some patients believed that TB is fatal, and this belief ultimately resulted in their depression. Engaging with the TB center was pronounced as emotional support and a reason for hope of recovery by many patients, and this support helped reduce their depression (Table-4). Patients and carers emphasized that social support for TB patients is a crucial key to their recovery. Due to TB, patients are already disturbed, and if they do not experience appropriate environments and care, their stress increases. They believed that communal settings, such as mosques and joint families, act as facilitators by providing emotional and financial support in times of need (Table-4). Likewise, the behaviour of doctors in TB centres is crucial, and patients often feel relieved only when the doctor reassures them about their disease and prognosis. (Table-3).

According to the Afghan participants, migration to Pakistan helped patients’ families to understand that TB is treatable. The common misconception about TB in Afghanistan is that it is incurable and fatal, and they used to separate the utensils of TB patients from those of other family members (Table-4). In Pakistan and Afghanistan, older people had more faith in the hakim’s (herbalist’s) treatment. Patients used to take their TB patients to a hakim whenever someone recommended a hakim for their patient’s illness. They visit them in the hope of achieving complete healing from many diseases (Table-3). Similarly, they believed that treatment for depression involves prayers and worship of Allah (God), and those who pray to Allah and are hopeful will completely recover (Table-4). Since patients in this region mostly give importance to religious practices and beliefs, health professionals also advise them to continue praying and engaging in religious remembrance to avoid depression (Table-2).

### TB and depression: the dual stigma

Healthcare providers believed that social stigma is inherently associated with TB and depression, and patients, due to stigma, are unable to discuss these conditions openly. With regards to stigma in Pashtun society, females are again the prime target; many young girls cannot tell their in-laws that they have been diagnosed with TB. Otherwise, their marriage could end or be delayed (Table-2). According to patients, TB is such a notorious disease that even family members start to distance themselves. Some say, “Don’t eat with us,” while others say, “Don’t cook food.” Observing this kind of behavior leads patients to feel depressed and ashamed. They begin to view themselves as a burden to society and their families, ultimately leading to profound feelings of loneliness (Table-3).

Participants of all groups agreed that if someone has TB or a mental health problem, they tend to hide it from others. If the community finds out that someone has TB or depression, the news will spread in a very negative way and create a spectacle. Both mental health problems and TB face a great deal of stigma in society (Table-3&4).

### Culturally preferred and acceptable therapy

When asked about counselling therapy, most patients and carers were aware of the routine counseling provided by healthcare providers at the TB facility regarding their medications and modifications to daily routines. They believed that such counseling by healthcare providers boosts their morale and can prevent them from experiencing depression. They preferred the notion of delivering psychosocial counseling therapy in the health facility rather than in other communal settings. They believed that half of the stress is reduced simply by talking to each other, as it helps clear the mind of stressful thoughts and lifts the mood. Patients highlighted the importance of involving family members in such counseling sessions (Table-3). In addition, some participants believed that counseling is more effective than pharmacological therapy for depressed patients as it improves the patients’ emotional strength and inspires courage in them (Table-3). Counseling sessions should include information about taking medications, addressing social issues, and the importance of maintaining a healthy diet.

Health professionals suggested that facility-based sessions are more effective in treating depression among TB patients. While some advocated for these sessions to be led by a psychologist, others felt trained staff could also provide effective psychotherapy. They emphasized the importance of offering incentives to families and patients attending these sessions (Table-2). Moreover, participants suggested that one-to-one sessions would be ideal because there is a stigma associated with both TB and mental health in society. Patients may not feel comfortable discussing certain matters in a group, which is why they should have separate sessions (Table-2). Some patients also suggested that if therapists could provide graphics-based booklets to take home, it would help keep their minds diverted (Table-4).

## Discussion

This is the first study to explore the comorbidity of TB and depression among the Pashtun ethnicity of Pakistan and Afghan nationals (living in Pakistan as refugees), eliciting the perceptions of healthcare providers, TB patients, and carers. The intersection between TB and mental health is influenced by cultural factors, especially in the context of LMICs. Thinking about their disease is inevitable among TB patients, often resulting in depression. This phenomenon is more pronounced among females who are already struggling with domestic problems and other social issues perpetuated by the joint family system, which is a common practice of the local culture. Moreover, the stigma related to both TB and depression is further exacerbating the two conditions, mainly by pushing the patients to social isolation and delaying them from seeking professional help. The positive role of family members and healthcare providers can be leveraged to help patients enhance their mental health.

Literature highlights a male dominance in Pashtun culture, with various discriminatory behaviors towards women. This gender discrimination is deeply rooted in the social fabric, where women cannot make decisions about their healthcare needs. They must obtain permission for their healthcare needs and must be accompanied by a male or an elder family member when visiting the healthcare facility ^22^. Our study yields similar findings: females are often accompanied by an attendee (carer) to the hospital and have no say in decisions regarding their health or the continuation of treatment. Cultural practices in other studies also report the prohibition of females to leave the house ‘unnecessarily’, including visits to health care facilities for conditions that are not life-threatening, because taking a woman out of the house is considered disrespectful ^23–25^. We found that female TB patients of Pashtun ethnicity face significant discrimination in society. Their health deteriorates due to TB, and at the same time, they lack support from their husbands and in-laws, causing distressing situations at home, leading to the onset of depression. Lack or diminished level of social support for female TB patients leading to depression has also been highlighted in other studies from LMICs like Bangladesh and Ethiopia ^26,27^.

Patients in the current study stressed the need for care and a supportive environment for recovery from depression, along with their illness: they were most comfortable with those healthcare providers who treated them with respect and listened to them. Effective doctor-patient interaction is an essential component in the delivery of healthcare ^28,29^. This positive communication between doctors and patients not only regulates emotions, enhances medical understanding, and identifies patient needs, but also leads to greater satisfaction, open information sharing, accurate diagnosis, and greater treatment adherence. ^29–31^. Spirituality and its association with disease and the healing process is a common belief among the Pashtun community, where illness is seen as God’s will and is considered directly or indirectly related to the sins of people. Many people believe that to recover, they must pray ^23,32,33^. These beliefs are consistent with our findings, and owing to the religious beliefs of participants, healthcare providers also advised them to pray and engage themselves in religious remembrance to avoid depression.

Our findings reveal significant stigma surrounding mental disorders and TB in Pakistan, causing reluctance to seek help due to fears of social ostracism and damage to reputation. Mental health issues are often viewed as a sign of personal weakness or a lack of faith. A study indicates that TB patients facing higher stigma are six times more likely to delay sputum testing, hindering their access to care and treatment completion ^34,35^. Similarly, TB-related stigma among patients has been reported to result in an 11 times increased likelihood of having depression in Ethiopia^36^. Stigma diminishes self-esteem, quality of life, and symptom disclosure, which hampers TB screening ^36,37^. Patients also tend to isolate due to social stigma, impacting their mental and physical health due to avoiding seeking help for their symptoms ^36,37^. The proven effects of stigma and poor social support on the cognitive and overall well-being of TB patients call for building effective social support mechanisms, which will encourage TB patients to seek timely assistance for psychological needs ^38^.

Our research emphasizes the effectiveness of culturally acceptable therapy through a patient-centered approach. Tailoring interventions to patients’ cultural backgrounds leads to better outcomes. A study of South Asian residents in Canada found that mental health services addressing family dynamics, cultural values, and socio-political factors are essential for reducing dropout rates in therapy for depression and anxiety ^39^. Limitations of the current study include that the findings are specific to two districts in KP, Pakistan, and may not apply to other regions within Pakistan or to other countries with different cultures. However, the findings are robust as we triangulated data from three different participant groups and two different nationalities. Although potential loss or misinterpretation of some nuances during translation may have happened, multiple IDIs and FGDs yielded deep insights into the personal and cultural contexts of TB and depression.

## Conclusion

This study examined the connection between tuberculosis (TB) and depression within the cultural and ethnic contexts of Pakistan, particularly among the Pashtun community and Afghan refugees. Cultural norms, social dynamics, and belief systems significantly impact the experiences of individuals facing these co-occurring health challenges, influenced by gender discrimination, financial hardships, strong (mis)beliefs, lack of social support, and stigma. To effectively address the medical and mental health needs of TB patients, comprehensive healthcare policies and psychosocial interventions that consider cultural contexts are essential. A holistic approach that enhances social support and doctor-patient communication is crucial for tackling the challenges of TB and its associated mental health burdens.

## Author statements

### Ethical approval

The study has received ethical approval from Keele University Ethical Review Panel (Ref: REC Project Reference 0599), Khyber Medical University Ethical Review Board (Ref: DIR/KMU-EB/CT/000999-2), and National Bioethics Committee Pakistan (Ref: 4-87/NBC-998/23/1517). Written permission was obtained from the Directorate of TB Control Program, KP-Pakistan. All ethical considerations were appropriately followed, including obtaining informed consent, ensuring participant anonymity, and maintaining confidentiality.

### Data availability statement

All relevant data is contained within the article: The original contributions presented in the study are included in the article; further inquiries can be directed to the corresponding author.

## Acknowledgements

We thank all the research team members for their tireless field work. Additionally, we would like to extend our gratitude to the TB patients and staff of the provincial TB control program in KP, Pakistan, for their continuous support.

